# Artificial Intelligence-Powered Precision Medicine for Cardiovascular Disease Prevention and Management

**DOI:** 10.1101/2025.03.23.25324474

**Authors:** Yudi Kurniawan Budi Susilo, Shamima Abdul Rahman, Kapil Amgain, Dewi Yuliana

## Abstract

Artificial intelligence (AI) is transforming precision medicine, particularly in cardiovascular disease prevention and management. This bibliometric analysis examines the research landscape from 2020 to 2024, focusing on AI’s role in improving diagnostics, personalizing treatment, and advancing predictive healthcare. Using the PRISMA framework, VOSviewer, Harzing’s Publish or Perish, and Excel, 137 articles from Scopus were systematically analyzed. The study reveals a significant surge in research activity, with 2024 marking a peak. Machine learning and deep learning are central to key advancements, enabling early detection and risk prediction. Contributions from leading institutions highlight the global and interdisciplinary nature of this field, with studies demonstrating AI’s potential to integrate complex datasets and deliver tailored therapies. While AI-driven innovations show promise, challenges such as ethical concerns and healthcare disparities remain. This analysis underscores AI’s transformative potential in precision medicine and identifies opportunities for equitable, collaborative advancements.

## 1.0 INTRODUCTION

The onset of the new millennium has witnessed a dramatic transformation in the management of cardiovascular diseases (CVDs), driven by the convergence of artificial intelligence (AI) techniques and the evolving paradigm of precision medicine [1], [2]. In recent years, the implementation of AI and machine learning (ML) has enriched the diagnostic and prognostic capabilities of traditional methodologies, facilitating early risk prediction and personalized therapeutic interventions in cardiovascular care [3], [4]. The integration of vast multi□modal datasets, such as genomic, clinical, lifestyle, and imaging data, has allowed for the development of sophisticated models that underpin P4 (predictive, preventive, personalized, and participatory) medicine, a framework that is gaining prominence in cardiovascular prevention and management [1], [5]. This innovative approach has not only enhanced the accuracy of disease classification but has also redefined the way clinicians approach treatment by tailoring strategies at an individual level [2], [6]. Furthermore, the fusion of AI with precision medicine represents a paradigm shift in clinical research aimed at revolutionizing patient outcomes through data-driven insights and dynamic risk stratification [4], [7].

## 2.0 LITERATURE REVIEW

The evolution of AI-powered precision medicine in the context of CVD prevention emerges as an interdisciplinary convergence of computer science, clinical medicine, and data analytics [2], [3]. Fundamentally, AI algorithms are now capable of processing extensive amounts of heterogenous data acquired from electronic health records, advanced imaging modalities, and wearable sensor devices, each contributing to a comprehensive risk profile [8], [9]. These advances resonate with the principles of precision medicine that prioritize individualized care by leveraging patient-specific information to optimize clinical decision-making [6], [5]. The contemporary literature highlights that the combination of high-throughput data acquisition and AI’s analytical capability opens new avenues for early detection and targeted intervention [1], [10]. Notably, the integration of explainable AI models has introduced transparency into the decision-making processes, thereby increasing the confidence of clinicians in adopting these novel diagnostic tools [11], [12].

Cardiovascular diseases remain the leading cause of death globally, prompting an urgent need for improved diagnostic and therapeutic methodologies [1], [13]. Traditional risk models based on clinical scoring systems have increasingly become insufficient in capturing the complexity and heterogeneity of CVDs [7], [14]. In contrast, AI-based approaches have demonstrated superior performance by detecting subtle patterns and biomarkers that often escape conventional detection methods [8], [15]. This shift towards predictive analytics is especially significant given that early and precise risk stratification can potentially mitigate the progression of cardiovascular events [2], [16]. As such, the current literature emphasizes the transition from generic treatment protocols to individualized management plans, accentuating the role of AI in realizing the full potential of precision cardiovascular medicine [17], [18].

The clinical applicability of AI in cardiovascular medicine is underpinned by its capacity to integrate vast clinical datasets into actionable insights [19], [17]. By harnessing deep learning and advanced ML algorithms, researchers have built models that predict the likelihood of cardiovascular incidents with high sensitivity and specificity [20], [21]. Moreover, these predictive models often incorporate real-world data, further enhancing their robustness and accuracy by reflecting the heterogeneous nature of patient populations [10], [22]. The proliferation of high-performance computing resources and the digitization of health records have accelerated the incorporation of AI tools into routine clinical practice [23], [24]. In summary, this transformative approach holds the promise of not only advancing diagnostic capabilities but also streamlining therapeutic interventions, thereby improving patient outcomes through precision-driven management [14], [25].

A significant component of precision medicine in cardiovascular care is the ability to personalize risk assessment and treatment strategies across diverse populations [2], [6]. The data amassed from multiple sources such as imaging, genomic sequencing, and wearable devices have laid the foundation for AI algorithms to delineate individual risk factors with exceptional specificity and granularity [26], [9]. Consequently, such technological advancements have spurred the development of targeted interventions designed to mitigate disease progression through early preventive measures [5], [13]. Clinically, these innovations facilitate a shift from reactive treatment modalities to proactive management of cardiovascular health, reflecting a critical advancement in public health strategy [1], [7]. Additionally, the iterative refinement of these AI-powered models through continuous learning and integration of new data is pivotal to sustaining their clinical relevance and accuracy [12], [17].

In recent systematic reviews and meta-analyses, precision medicine has been recognized as a transformative approach that addresses the inherent variability in patient responses to treatment [7], [17]. The ability to harness AI for aggregating and analyzing large-scale patient data is central to this progress, providing insights into disease mechanisms and therapeutic response [2], [3]. With continuous improvements in computing technology, sophisticated algorithms are now standard in translating clinical and multi-omics data into refined diagnostic tools [4], [12]. These advances represent a convergence point between computational innovations and clinical necessity, thereby reinforcing the relevance of AI-enabled precision medicine in cardiovascular disease management [8], [15]. The holistic synthesis of diverse data types through AI fosters a more accurate and individualized prediction model that emphasizes prevention over treatment [5], [13].

## 3.0 METHODS

This study adopts a systematic bibliometric analysis to explore the research landscape surrounding artificial intelligence, precision medicine, cardiovascular disease, and prevention. A structured approach was employed, integrating the PRISMA framework for screening and selection, along with tools such as VOSviewer, Harzing’s Publish or Perish, and Microsoft Excel for data extraction, analysis, and visualization.

The data for this study was sourced from the Scopus database, chosen for its extensive and high-quality coverage of peer-reviewed journals. The search strategy involved a carefully constructed keyword query encompassing terms like “artificial intelligence,” “precision medicine,” “cardiovascular disease,” and “prevention.” The search parameters were restricted to journal articles published in English between 2020 and 2024. This initial search yielded 426 records, which were subjected to further screening.

The PRISMA framework was employed to ensure a transparent and systematic screening process. Duplicate records were removed, and articles that fell outside the scope of the study were excluded. The inclusion criteria focused on publications that addressed the intersection of artificial intelligence and healthcare, particularly those exploring precision medicine and cardiovascular disease. Following the screening process, 137 records were identified as relevant and included for bibliometric analysis.

The analysis relied on several specialized tools to extract meaningful insights from the data. VOSviewer was used to visualize and analyze bibliometric networks, including keyword co-occurrence maps and collaborative relationships between countries. These visualizations helped to identify key research themes, clusters, and partnerships. Harzing’s Publish or Perish was utilized to calculate critical citation metrics such as the h-index, g-index, and citations per paper, providing a quantitative assessment of the research impact. Microsoft Excel was employed for data preprocessing, including filtering, categorization, and performing quantitative analyses to uncover trends in publication activity, journal contributions, and author productivity.

The results of this analysis offered valuable insights into the research dynamics of artificial intelligence-driven healthcare advancements. Key findings included trends in publication and citation activity, contributions of leading authors and institutions, and thematic clusters within the field. The combination of bibliometric tools and a systematic approach ensured the findings were robust, reproducible, and reflective of the current state of research. This methodology provides a comprehensive overview of the topic, offering a foundation for identifying gaps and opportunities for future research directions.

## 4.0 RESULTS

### 4.1 Analysis by year

The analysis of document publications over the past five years reveals a steady growth in research activity, culminating in a significant surge in 2024. With 50 documents contributing 36.50% of the total, 2024 stands as the most prolific year, marking a peak in research productivity. This sharp rise could be attributed to advancements in the field, increased global focus, or a higher allocation of resources towards the subject. The significant jump from 2023 to 2024 highlights the field’s growing importance, underscoring its relevance in addressing emerging challenges or opportunities.

Between 2020 and 2022, a consistent upward trend is observed, with 13 documents (9.49%) in 2020, 18 documents (13.14%) in 2021, and 26 documents (18.98%) in 2022. This gradual increase reflects a solid foundation of interest and effort that likely set the stage for the more pronounced growth in later years. The relative scarcity of publications in 2020 might be explained by global disruptions or an earlier stage of interest in the topic. By 2022, however, the momentum had clearly gained traction, leading to the notable rise seen in subsequent years.

The substantial increase in publications over the five-year period demonstrates the rapid evolution and growing significance of this research area. The fourfold growth from 2020 to 2024 not only reflects the field’s expanding scope but also highlights its potential for continued innovation. The trends observed suggest a strong trajectory of interest, making it worthwhile to investigate the underlying factors driving this growth, particularly in the peak year of 2024, to better understand the dynamics shaping this area of study.

### 4.2 Documents by source title

The distribution of documents across various source titles highlights the diverse outlets contributing to the research in this field. Among the journals, the Journal of Personalized Medicine leads with 7 documents, representing 5.15% of the total. This dominance underscores the journal’s significant focus on cutting-edge developments in personalized approaches to healthcare, aligning with the broader theme of precision medicine and artificial intelligence. The Epma Journal follows closely with 6 documents (4.41%), further emphasizing the importance of predictive, preventive, and personalized medicine in this research domain.

Other journals, such as Frontiers in Cardiovascular Medicine and the Journal of the American Heart Association, each contribute 5 documents, accounting for 3.68% of the total. These journals are highly regarded for their focus on advancements in cardiovascular research and related innovations, which are pivotal for understanding the applications of artificial intelligence in this domain. A group of journals, including Circulation, the European Heart Journal, IEEE Access, Journal of Clinical Medicine, and PLOS ONE, each contribute 3 documents (2.21%). This shows a balanced representation of multidisciplinary research, ranging from clinical studies to technological innovations.

At the lower end of the spectrum, the American Heart Journal Plus Cardiology Research and Practice contributes 2 documents (1.47%), and the IEEE Transactions on Consumer Electronics features 1 document (0.74%). While these contributions are smaller in number, they reflect niche areas of study or emerging intersections between technology and medicine. Collectively, the distribution of publications across these source titles demonstrates the interdisciplinary nature of the research, with contributions from clinical, technological, and personalized medicine-focused journals.

### 4.3 Document by affiliation

The distribution of documents by institutional affiliation reveals significant contributions from several leading organizations. Topping the list is *Brigham and Women’s Hospital*, with 7 documents accounting for 2.03% of the total publications. This highlights the institution’s prominent role in advancing research, particularly in the areas of medicine and healthcare innovation. Its strong representation underscores its commitment to cutting-edge studies and interdisciplinary collaboration.

*Harvard Medical School* and *Universität Bonn* both contribute 6 documents, each representing 1.74% of the total. These institutions are renowned for their excellence in research and academic rigor, suggesting their involvement in pivotal studies that drive the field forward. Their presence in the list reflects their sustained focus on impactful and high-quality research outputs.

Several other affiliations, including the *Ministry of Education of the People’s Republic of China, Mayo Clinic, Stanford University, University of California, Los Angeles*, and *Universitätsklinikum Bonn*, each contributed 5 documents, making up 1.45% of the total. These contributions demonstrate the global and collaborative nature of research in this domain, with participation from institutions across diverse geographical regions and specializations. Their involvement highlights the importance of combining clinical, technological, and educational expertise to address complex challenges.

Finally, *Johns Hopkins University School of Medicine* and the *David Geffen School of Medicine at UCLA* each contributed 4 documents (1.16%). These institutions, though contributing slightly fewer documents, are still influential players in the field. Their inclusion reflects the diversity of expertise and the broad institutional base supporting advancements in the domain. The analysis shows that institutions with strong clinical, academic, and research foundations play a pivotal role in driving innovation and expanding knowledge. The wide representation of affiliations reflects the collaborative and interdisciplinary nature of the research, involving key players from across the globe.

### 4.4 Document by subject area

The analysis of documents by subject area highlights the interdisciplinary nature of the research field, with a strong emphasis on medicine. With 102 documents accounting for 49.76% of the total, medicine is the leading subject area, underscoring its pivotal role in driving advancements and applications in this domain. The dominance of medicine reflects the centrality of healthcare-related topics, particularly in the integration of technology and innovation to address clinical challenges.

Other significant contributors include *Pharmacology, Toxicology, and Pharmaceutics* with 17 documents (8.29%) and *Biochemistry, Genetics, and Molecular Biology* with 16 documents (7.80%). These areas emphasize the importance of drug development, molecular research, and genetic studies, which are foundational to personalized medicine and other precision healthcare initiatives. Their contributions highlight the growing intersection of life sciences and technology in advancing healthcare solutions.

*Computer Science* also plays a notable role with 15 documents (7.32%), showcasing the critical involvement of technological disciplines in supporting medical and scientific research. The contributions of *Engineering* (11 documents, 5.37%) and *Health Professions* (9 documents, 4.39%) further reinforce the collaborative nature of this field, combining computational expertise, engineering solutions, and healthcare delivery to innovate in diagnosis, treatment, and management.

Other areas, including *Multidisciplinary* studies (8 documents, 3.90%), *Materials Science* (5 documents, 2.44%), *Neuroscience* (5 documents, 2.44%), and *Agricultural and Biological Sciences* (4 documents, 1.95%), demonstrate the breadth of research applications. These fields contribute unique perspectives and methodologies, enriching the overall research ecosystem and fostering innovation across domains.

The distribution of documents across these subject areas highlights the importance of collaboration between medicine, technology, and the life sciences. This interdisciplinary approach is essential for addressing complex healthcare challenges and underscores the diverse contributions that drive progress in the field.

### 4.5 Keyword analysis

The keyword analysis reveals critical themes and priorities in the research field, highlighting the central role of machine learning and artificial intelligence (AI) in advancing healthcare innovation. *Machine learning* emerges as the most frequently occurring keyword, with 45 mentions, reflecting its pivotal role as a core technology in healthcare applications. Closely following, *artificial intelligence* with 24 occurrences underscores its broader significance as a foundation for various advancements, from diagnostics to treatment optimization. Together, these technologies serve as the backbone for implementing precision and personalized medicine.

The prominence of keywords such as *precision medicine* (23 occurrences) and *personalized medicine* (17 occurrences) emphasizes the increasing shift toward tailoring healthcare interventions to individual patient needs. This trend reflects the transformative potential of AI-driven approaches in enhancing healthcare outcomes by leveraging data-driven insights. The close connection between these terms and *machine learning* in the network highlights their interdependence, showcasing how computational advancements are revolutionizing the delivery of medical care.

The network also reveals a strong focus on cardiovascular health, with keywords such as *cardiovascular diseases* (10 occurrences), *heart failure* (8 occurrences), and *coronary artery disease* (7 occurrences) occupying prominent positions. These terms signify the ongoing efforts to address one of the leading causes of mortality globally using AI and machine learning. Additionally, the inclusion of *COVID-19* (7 occurrences) as a keyword reflects the responsiveness of the research community to pressing global challenges. It demonstrates how AI technologies have been rapidly adapted to combat emerging health crises, particularly in diagnostics and management.

The interconnectedness within the keyword network illustrates the interdisciplinary nature of this research field. Terms like *deep learning* (7 occurrences) and *healthcare* (6 occurrences) show the integration of advanced methodologies and broader systemic considerations. This interconnected framework underscores the collaborative effort between medical, technological, and scientific disciplines to innovate and solve complex healthcare challenges.

In conclusion, the keyword analysis and network insights highlight a dynamic research landscape driven by the convergence of AI and healthcare. The strong emphasis on machine learning, precision medicine, and cardiovascular health demonstrates the field’s focus on addressing critical healthcare needs through innovative, data-driven solutions. Additionally, the inclusion of emerging topics such as COVID-19 showcases the adaptability of AI technologies in responding to global challenges, paving the way for continued progress and transformative impact in the field of medicine.

### 4.6 Analysis of the Top 10 Documents by Citation

The table of the top 10 most-cited documents provides a clear snapshot of influential research in the field. The highest-cited paper, authored by *I. Landi et al*., received 107 citations for its contribution to the application of deep learning in electronic health records, emphasizing the role of artificial intelligence in patient stratification. Published in *Nature Research* in 2020, the paper underscores the value of advanced computational techniques in healthcare innovation.

*W. Wang et al*. secured the second position with 95 citations for their joint position paper discussing suboptimal health and personalized medicine, highlighting the integration of predictive techniques for better health outcomes. Similarly, *O. Golubnitschaja et al*. contributed significantly with 80 citations for their 2021 *EPMA Position Paper*, focusing on health risks associated with BMI classifications, which has implications for precision medicine.

Other key contributions include *W. DeGroat et al*.’s work (76 citations) on discovering biomarkers for cardiovascular diseases using machine learning, published in *Nature Research* in 2024, and *R*.*M. Hoogeveen et al*.’s study (75 citations) on plasma proteomics for cardiovascular risk prediction, published in *Oxford University Press* in 2020. These papers exemplify the increasing importance of machine learning and omics-based approaches in medical research.

The list further includes research by *O. Strianese et al*. (67 citations) on genomic approaches for cardiovascular and neurodegenerative diseases and *G*.*Y*.*H. Lip et al*. (66 citations) on machine learning models for stroke risk prediction. These papers demonstrate the diversity of applications for AI and data analytics in addressing critical healthcare challenges.

Finally, contributions from *C. Napoli et al*. (58 citations) on epigenetic factors in cardiovascular risk and *G. Adami et al*. (51 citations) on osteoporosis predictions provide insights into the expanding domains of precision medicine. *C*.*B. Collin et al*. (44 citations) on computational models in personalized medicine concludes the list, underscoring the importance of integrating data for model validation.

### Conclusion

The top 10 most-cited papers illustrate the profound impact of machine learning, AI, and precision medicine on modern healthcare. With contributions spanning from electronic health records to cardiovascular research and epigenetics, these papers reflect the interdisciplinary and innovative nature of the field. The focus on predictive, preventive, and personalized approaches underscores the ongoing transformation of healthcare driven by advanced technologies. These studies not only highlight current achievements but also set the stage for future research and clinical applications.

## 5.0 DISCUSSION

The findings of this bibliometric analysis highlight the profound impact and diverse applications of AI in cardiovascular precision medicine. Over the analyzed period (2020–2024), there has been a marked increase in research activity, culminating in a significant surge in publications in 2024. This growth reflects the expanding adoption of AI-driven methodologies in addressing the complexities of cardiovascular diseases. Machine learning and deep learning emerge as central themes, underpinning advancements in early detection, risk prediction, and treatment optimization.

Key studies, such as those by DeGroat et al. (2024), demonstrate the efficacy of AI in identifying biomarkers predictive of cardiovascular disease, achieving high levels of diagnostic accuracy. This aligns with broader trends of integrating omics-based data into clinical practice, facilitating precision medicine. Furthermore, the collaborative nature of research in this field is evident from the strong co-authorship networks and the contributions of leading institutions like Brigham and Women’s Hospital and Harvard Medical School. These collaborations have enabled the translation of AI technologies from experimental frameworks to practical clinical applications.

However, challenges remain in ensuring the equitable deployment of AI-driven healthcare solutions. Ethical considerations, such as data privacy, bias in algorithms, and accessibility, must be addressed to maximize the societal impact of these advancements. Additionally, the variability in research output across regions underscores the need for increased global collaboration to bridge gaps in technology adoption and healthcare delivery. Despite these challenges, the trajectory of research suggests a promising future for AI in transforming cardiovascular precision medicine, with potential applications extending to other domains of healthcare.

## 6.0 CONCLUSION

This study underscores the transformative potential of artificial intelligence in cardiovascular precision medicine, particularly in enhancing diagnostic accuracy, personalizing treatment, and preventing disease progression. The bibliometric analysis highlights the significant growth in research activity, driven by technological advancements and increasing global collaboration. Key contributions from leading researchers and institutions have propelled the integration of AI into clinical practice, addressing critical healthcare challenges.

The findings emphasize the interdisciplinary nature of this research field, bridging medicine, computer science, and engineering. While significant progress has been made, challenges related to ethical considerations, data accessibility, and regional disparities must be addressed to fully realize the potential of AI in healthcare. Moving forward, fostering international collaboration and focusing on patient-centered innovations will be crucial in leveraging AI to transform cardiovascular medicine and beyond. By continuing to refine and expand the applications of AI, researchers and practitioners can usher in a new era of predictive, preventive, and personalized healthcare.

## Data Availability

All data produced are available online at:
1. https://doi.org/10.17632/hhv937v7pv.1
2. https://www.kaggle.com/datasets/yudi299/artificial-intelligence-powered-precision-medicine

https://doi.org/10.17632/hhv937v7pv.1

https://www.kaggle.com/datasets/yudi299/artificial-intelligence-powered-precision-medicine

## Dataset reference

Budi Susilo, Yudi Kurniawan (2025), “Artificial Intelligence-Powered Precision Medicine for Cardi-ovascular Disease Prevention and Management”, Mendeley Data, V1, doi: 10.17632/hhv937v7pv.1

---

**Figure 1.**
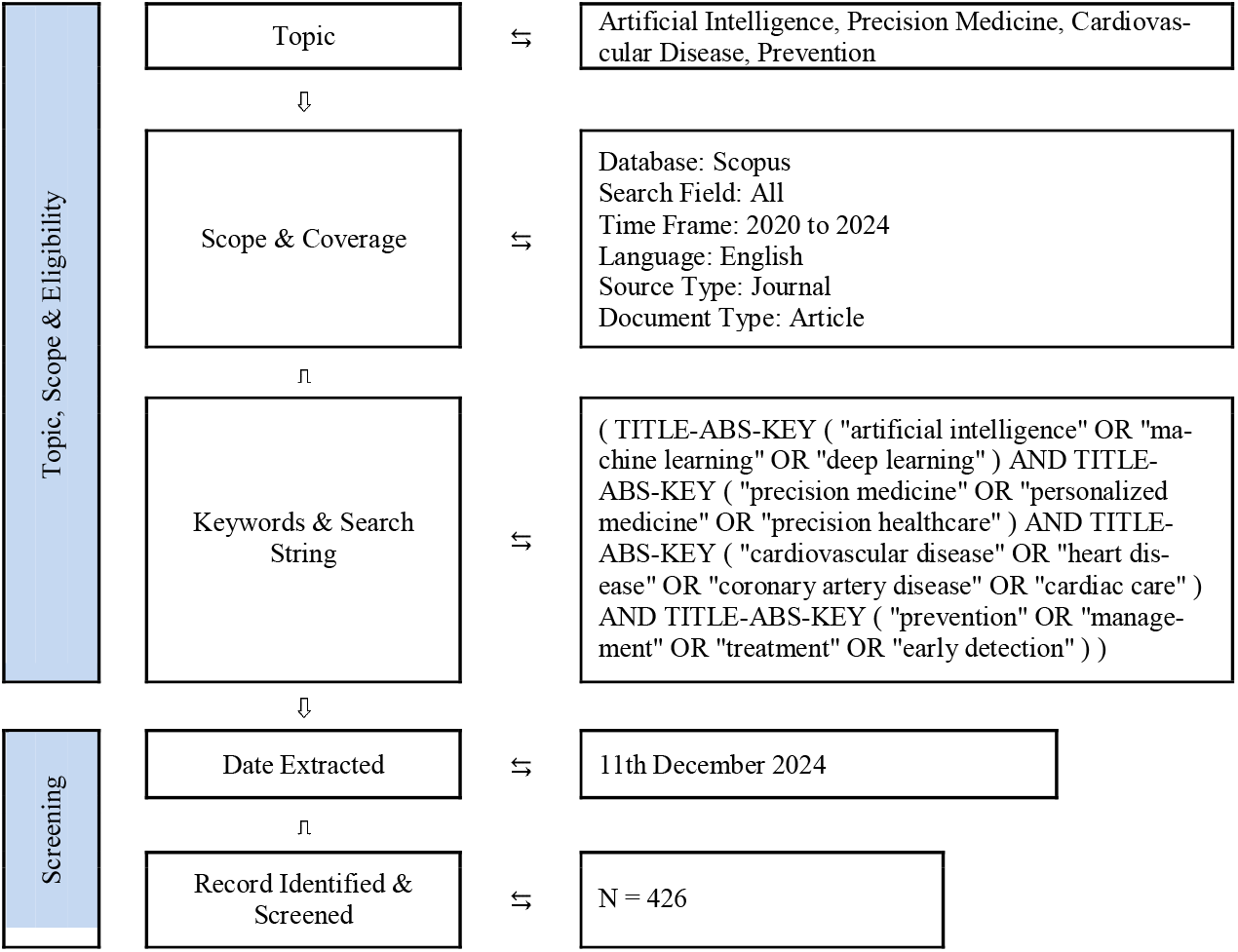
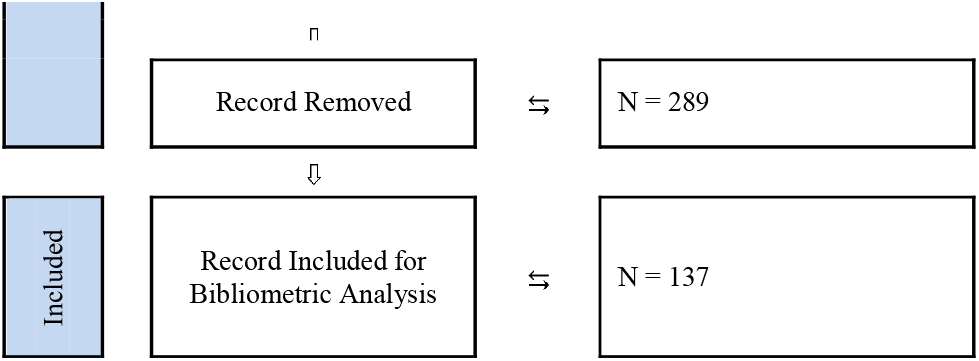
Flow diagram of the search strategy to explore the research landscape surrounding artificial intelligence, precision medicine, cardiovascular disease, and prevention

**Figure 2.**
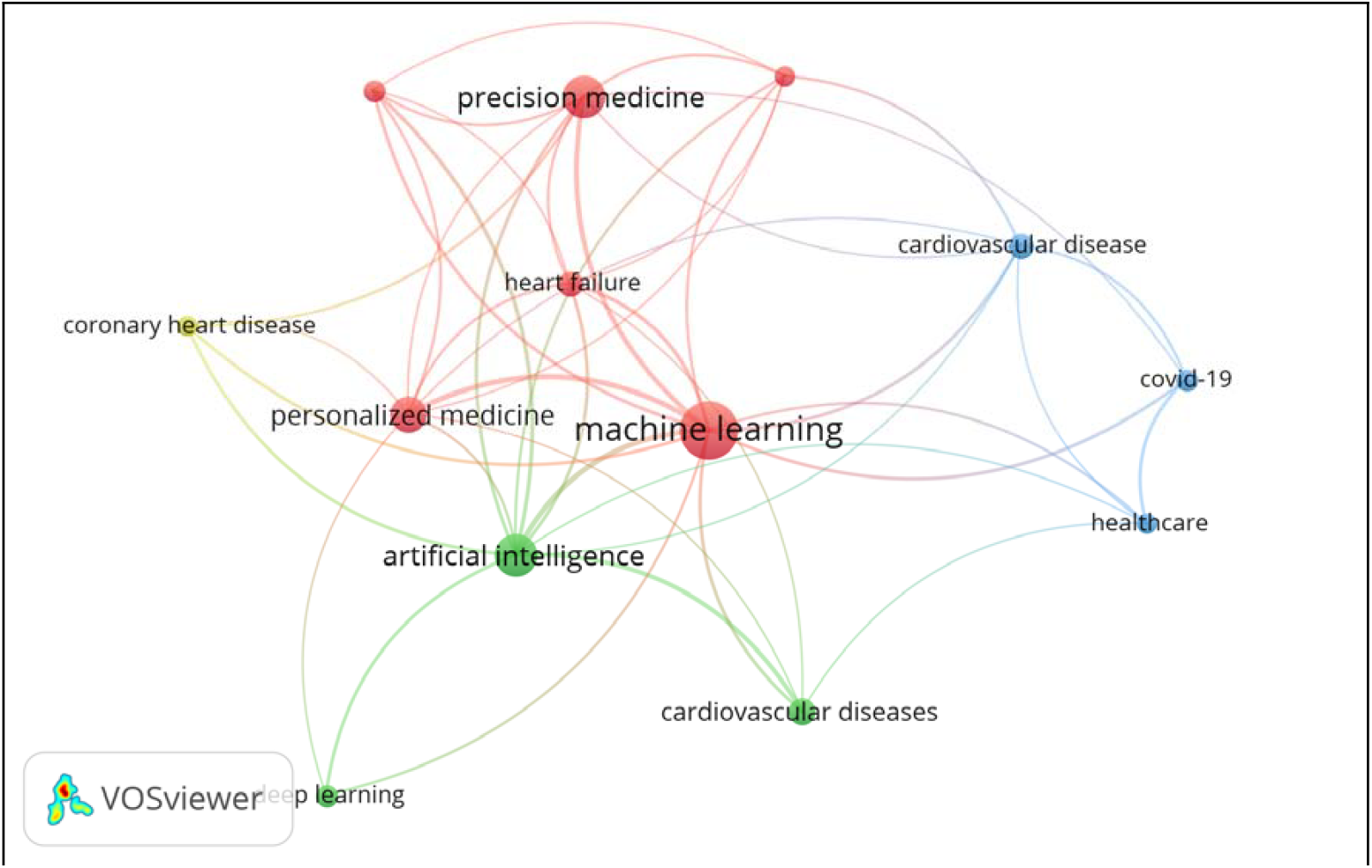
Network visualization of bibliographic coupling links for keyword co-occurrence

**Table 1.** Documents by year over the past five years (2020 to 2024)

| Year | Documents | Percentage (%) |
| --- | --- | --- |
| 2024 | 50 | 36.50 |
| 2023 | 30 | 21.90 |
| 2022 | 26 | 18.98 |
| 2021 | 18 | 13.14 |
| 2020 | 13 | 9.49 |

**Table 2.** Documents by source title contributing to the research field

| Source title | Documents | Percentage (%) |
| --- | --- | --- |
| Journal Of Personalized Medicine | 7 | 5.15 |
| Epma Journal | 6 | 4.41 |
| Frontiers In Cardiovascular Medicine | 5 | 3.68 |
| Journal Of the American Heart Association | 5 | 3.68 |
| Circulation | 3 | 2.21 |
| European Heart Journal | 3 | 2.21 |
| IEEE Access | 3 | 2.21 |
| Journal Of Clinical Medicine | 3 | 2.21 |
| Plos One | 3 | 2.21 |
| American Heart Journal Plus Cardiology Research and Practice | 2 | 1.47 |
| IEEE Transactions on Consumer Electronics | 1 | 0.74 |

**Table 3.** Document by affiliation significant contributions from several leading organizations

| Affiliation | Documents | Percentage (%) |
| --- | --- | --- |
| Brigham and Women's Hospital | 7 | 2.03 |
| Harvard Medical School | 6 | 1.74 |
| Universität Bonn | 6 | 1.74 |
| Ministry of Education of the People's Republic of China | 5 | 1.45 |
| Mayo Clinic | 5 | 1.45 |
| Stanford University | 5 | 1.45 |
| University of California, Los Angeles | 5 | 1.45 |
| Universitätsklinikum Bonn | 5 | 1.45 |
| Johns Hopkins University School of Medicine | 4 | 1.16 |
| David Geffen School of Medicine at UCLA | 4 | 1.16 |

**Table 4.** Document by subject area interdisciplinary nature of the research field

| Subject area | Documents | Percentage (%) |
| --- | --- | --- |
| Medicine | 102 | 49.76 |
| Pharmacology, Toxicology and Pharmaceutics | 17 | 8.29 |
| Biochemistry, Genetics and Molecular Biology | 16 | 7.80 |
| Computer Science | 15 | 7.32 |
| Engineering | 11 | 5.37 |
| Health Professions | 9 | 4.39 |
| Multidisciplinary | 8 | 3.90 |
| Materials Science | 5 | 2.44 |
| Neuroscience | 5 | 2.44 |
| Agricultural and Biological Sciences | 4 | 1.95 |

**Table 5.**
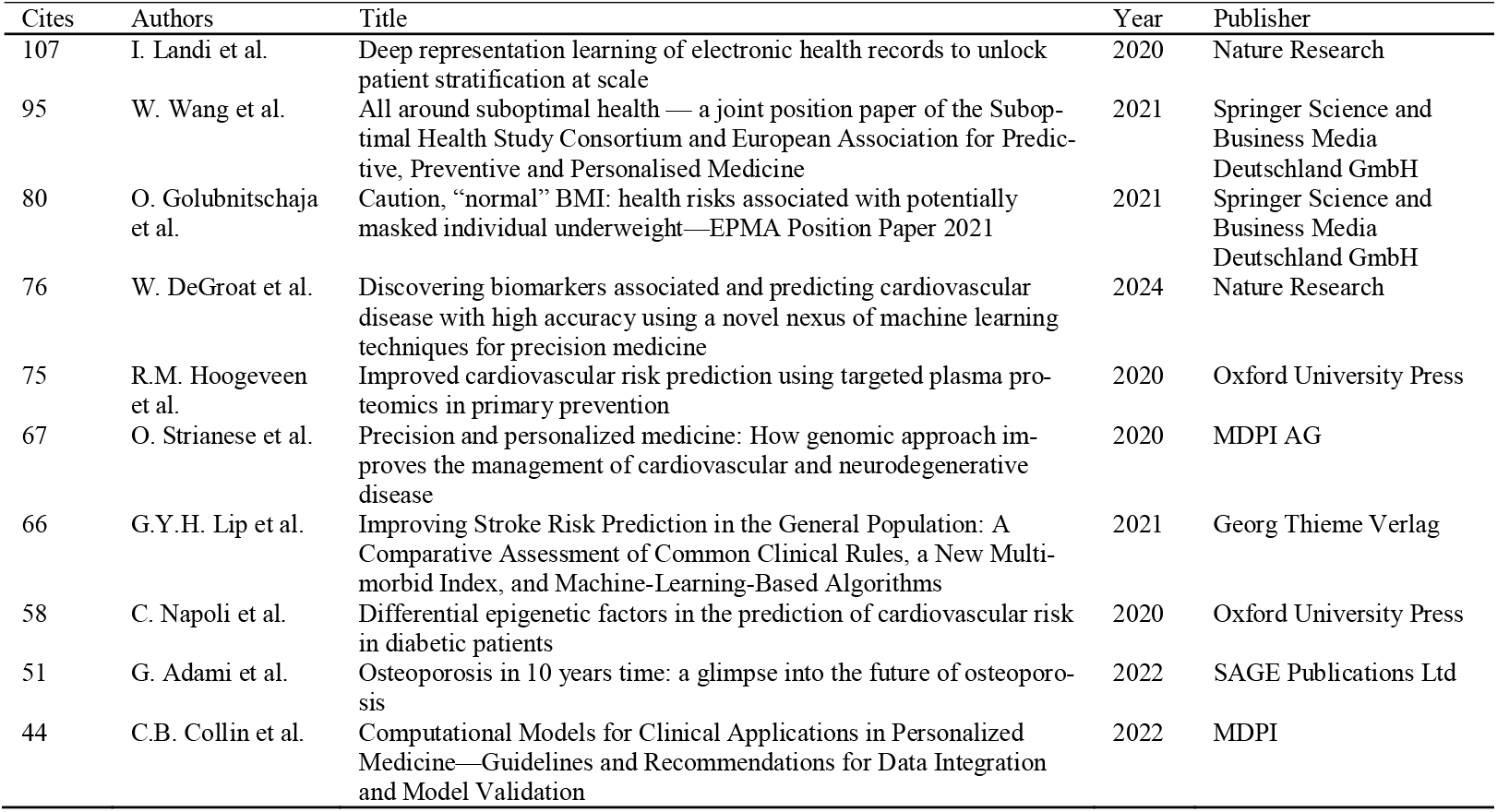
Top 10 Document by citation of influential research in the field

